# Excess Deaths associated with the COVID-19 Pandemic in Ukraine in 2020

**DOI:** 10.1101/2021.09.28.21264266

**Authors:** Neil K. Mehta, Ihor Honchar, Olena Doroshenko, Igor Brovchenko, Khrystyna Pak, Maria Danyuk, Pavlo Polikarchuk

**Author notes:** Corresponding author: Dr. Neil K. Mehta, The University of Texas Medical Branch, Maurice Ewing Hall/ Suite 1.128, Galveston, TX 77550.

## Abstract

COVID-19 related mortality has been understudied in Ukraine. As part of a World Bank project, we estimated excess mortality in Ukraine during 2020. Data on all deaths registered in government-controlled Ukraine from 2016-2020 (N=2,946,505) were utilized. We predicted deaths in 2020 by five-year age groups, sex, and month and calculated the number of deaths that deviated from expected levels (excess deaths). We compared excess deaths with the number of recorded COVID-19 deaths on death certificates and with published estimates for 30 European countries. We estimated 38,095 excess deaths in 2020 (6% of all deaths). Death rates were above expected levels in February and from June-December and lower in January and March-May. From June-December, we estimated 52,124 excess deaths with a peak in November (16,891 deaths). COVID-19 recorded deaths were approximately one-third of excess deaths in June-December (18,959 vs. 52,124). Higher than expected mortality was detected for all age groups 40-44 years and above and for those ages 0-4, 15-19, and 20-24. Ukraine’s excess mortality was about average compared to 30 other European countries. Excess deaths may be attributed directly to SARS-COV2 infection or indirectly to death causes associated with social and economic upheavals resulting in from the pandemic. Lower than expected mortality during the early part of 2020 is consistent with low influenza activity and reductions in deaths from restricted movement. Further studies are required to examine the causes of death that have contributed to positive excess mortality, particularly among younger aged groups.

**Key Messages:**

- Ukraine has experienced sizeable changes in its recent demography and the impact of the COVID-19 pandemic on the country’s aggregate mortality patterns is understudied
- Based on recent death trends, we found that Ukraine experienced lower than expected mortality during the early part of 2020 and consistently higher than expected mortality from June-December with peak levels occurring in November
- Positive excess mortality was observed for all age groups beginning at ages 40-44 as well as some younger age groups.

## Introduction

As the COVID-19 pandemic enters its second year, nations continue to grapple with its impact on nearly all facets of society. One critical indicator of the pandemic’s effect is its toll on national-level death rates. Early estimates indicate that the pandemic’s effect in this regard is sizeable with many countries experiencing death rates that are more than 40% higher than usual during peak months of the epidemic [1–3]. Death rates in the European Union (EU) as a whole, for example, were 25% higher in April 2020 than they were in the average of the same month in the previous four years [2]. In this paper, we provide estimates of excess deaths in 2020 for Ukraine based on recent death trends.

Like many European countries, Ukraine registered its first case of COVID-19 in March (3^rd^ March 2020). The country implemented its first quarantine on the 12^th^ March [4]. Despite this early intervention, the number of confirmed daily cases grew rapidly to approximately 100 per day at the beginning of April [5]. Stricter measures were instituted on 6^th^ April 2020, which included the closing of schools, shopping malls, and restrictions on travel [4]. These strict restrictions were in place until 22^nd^ May when the government eased some restrictions and moved to a policy of “adaptive quarantine”. While the growth rate of daily cases slowed after the stricter measures were instituted daily cases continued to grow reaching a peak of approximately 14,000 daily cases at the end of November. By the end of December, Ukraine was recording approximately 8,100 daily cases.

To date, the impact of the pandemic on Ukrainian mortality patterns remains poorly understood. While estimates of 2020 “excess deaths” for Ukraine are available online in international databases these databases do not provide detailed information on age and sex groups [2,3]. International databases also utilize crude methods for estimating excess mortality, relying on “average” death rates in the previous years. The assumption from these models is that the number of deaths is stable on a year-to-year basis. While the assumption may be valid for countries whose population size and age structure are relatively stable, it is not valid for countries experiencing demographic changes such as fluctuating migration levels, rapid aging, changing mortality levels, or population decline. Ukraine has experienced each of these processes over the last five years and we identify the misspecifications that can arise from reliance on the “average” method.

We estimated excess deaths in Ukraine for 2020 and compare these estimates with the number of deaths with COVID-19 reported on the death certificate. While older individuals face a substantial risk of death if infected by the novel coronavirus [6], the magnitude of the effect of the pandemic on younger aged mortality is less clear. However, there are reasons to believe that younger individuals are also experiencing significant excess mortality, which is supported by a growing evidence base [7,8]. Working-aged individuals may face higher risks of infection compared to other age groups because of workplace exposures. They may also be more susceptible to the economic disruptions caused by the pandemic compared to those who are retired. Adolescents and young adults may also experience excess mortality from risky behaviors and the negative effects of social isolation (e.g., suicides). Therefore, we estimated excess deaths for five-year age groups beginning at birth in addition to providing aggregate national estimates.

## Methods

### Data

We compiled deaths by five year groups (0-4, 5-9, …, 80-84) and for 85+, sex, and calendar month for each year during 2015-209 using data from the Ministry of Justice of Ukraine. We additionally compiled data on monthly deaths with COVID-19 on the death certificate obtained from the Public Health Center of Ukraine (PHCU), which we term COVID-19 coded deaths. Data on total and COVID-19 coded deaths were current as of February 2021. In addition to collecting data on deaths, which were used in our model, we separately reconstructed the age- and sex distribution of the Ukrainian population for 2016-2020 to provide additional context of the population of Ukraine. This component of the project was done with support from the Swiss Agency for Development and Cooperation. Ukraine has not administered a census of population for nearly 20 years. We began with the official population counts available for December 1, 2019 [9] and projected the population forward by age and sex to January 1, 2020. We then reconstructed the age and sex distribution of the population on January 1 in each year during 2016-2019. The projection and reconstruction of the population was based on a cohort component method accounting for natural change (births and deaths) and net migration. Population estimates were developed by sex and for each single year of age. We did not reconstruct the population data for 2015 because the data by age were not reliable because of the conflict. Data for the population reconstruction were obtained from the State Border Guard Service of Ukraine, State Statistics Service of Ukraine, Ministry of Health, PHCU. The estimates that we produced are pertinent to the Ukrainian controlled areas of the region and will differ from the World Population Prospects published by the United Nations as those data are pertinent to the entirety of Ukraine.

To calculate excess deaths in 2020 we implemented a model-based approach leveraging recent trends in mortality. Data from 2015-2019 by calendar month were used. We first estimated the monthly seasonal index, defined as the ratio of actual deaths in a month to the average monthly deaths over that calendar year. Next, we estimated a linear regression model with monthly deaths as the outcome: *Y*_*t*_ = *β*_0_ + *β*_1_ * *Month _t_*; where *t* indexes the calendar month with *t=*1 denoting January 2015 and *t*=60 denoting December 2019. We then predicted deaths for each month in 2020 (months 61 to 72). To arrive at our predicted deaths, each of the predicted monthly deaths for 2020 was multiplied by the 2015-2019 average seasonal index for that month. We calculated monthly 2020 excess deaths by subtracting observed monthly deaths from the predicted number of monthly deaths. We compared our estimates of excess deaths to the number COVID-19 coded deaths.

A strength of our approach is that it averages out seasonal fluctuations in mortality, which vary considerably from year to year primarily due to influenza and secondarily weather patterns. We validated our model by predicting monthly deaths for 2015-2019 based on the model and comparing them to actual monthly deaths (Appendix 1). The predicted and actual monthly deaths differed by an average of 1.36% suggesting that the model performed well. The mean standard error of the model was 2.1%. We present 95% confidence intervals for the monthly estimates of 2020 excess mortality.

We further applied our procedure to five-year age groups (0-4 to 80-84) and to ages 85+. Those with unknown age were excluded from the age-specific predictions (less than 0.25% of deaths annually did not have a recorded age). Lastly, we compared our estimates of Ukrainian monthly excess deaths with that published by Eurostat for European Union (EU) countries [2].

## Results

Table 1 provides population characteristics of the government-controlled Ukrainian population for 2016-2019. Between 2016 and 2019 the population declined from just under 38.2 million people to 37.3 million. Over this period the percentage over aged 65 years increased from 17% to 18%. The death rate (deaths per 100,000 persons) was highest in 2020 and lowest in 2017. There were 19,718 COVID-19 coded deaths in 2020. The COVID-19 coded death rate was 53 deaths per 100,000 persons, which indicates that approximately 3.2% of all deaths in 2020 were COVID-19 coded.

**Table 1.**
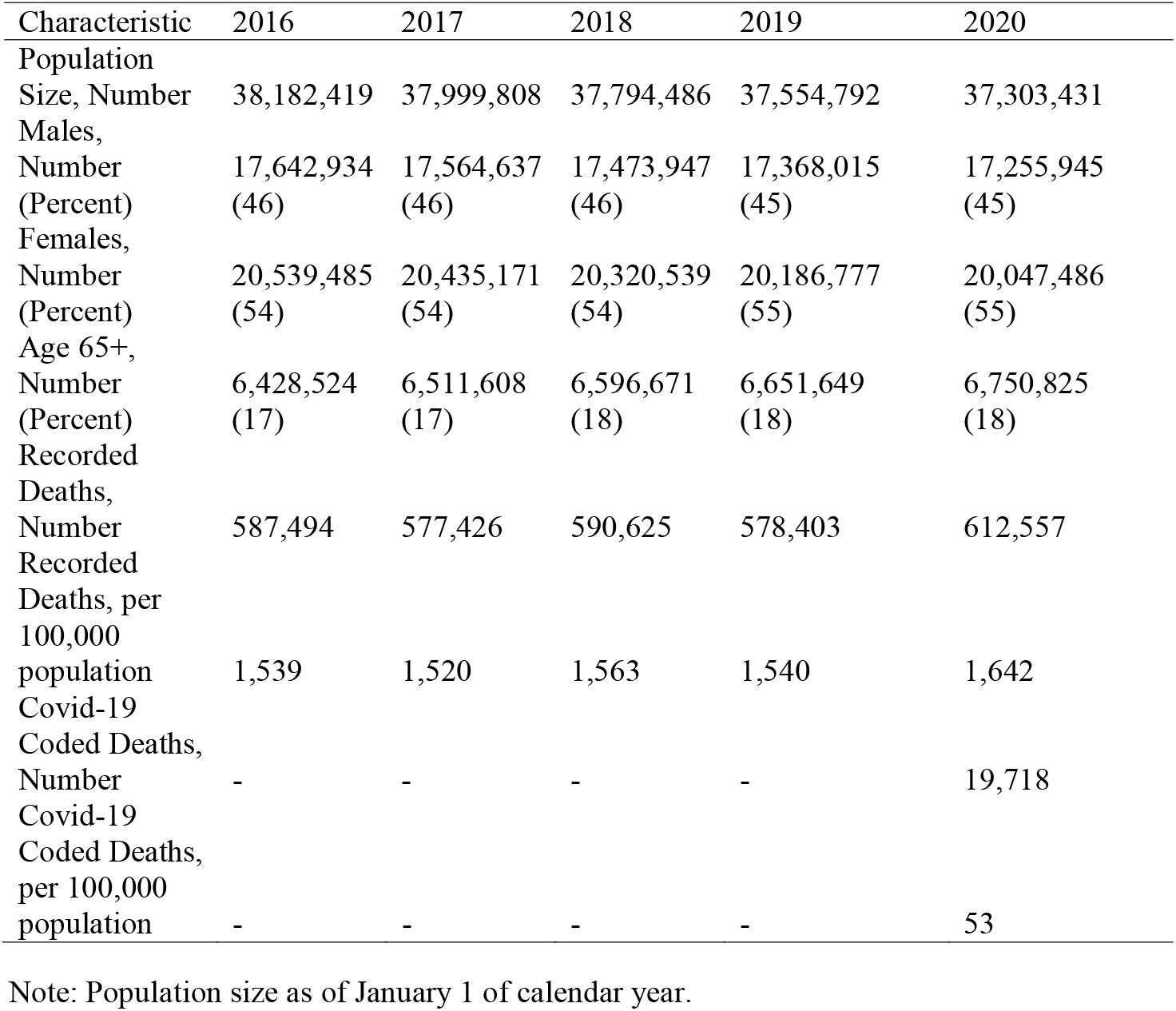
Descriptive characteristics of the government-controlled Ukrainian population, all ages

Figure 1 displays mortality and the seasonality index by year and calendar month. The figure highlights both the seasonal pattern of mortality and the distinct pattern of mortality for 2020 compared to the earlier years. In each year, mortality was highest in the fall and winter months and lowest in late summer. Mortality in 2020 was equal to or lower than the earlier years until approximately the middle of May after which rates were higher in 2020 than in each of the previous years (Figure 1a). Note that February 2020 was a leap year with an extra day, thus the 2020 death rate for this month encompasses slightly more exposure than in the previous years. The seasonality index for 2020 was approximately 1.10 in October and then steadily increased thereafter reaching a peak of 1.25 in November (Figure 1b). Earlier years also experienced a peak seasonality index of close to 1.25, however, this usually occurred at the beginning of the year and was short-lived.

**Figure 1.**
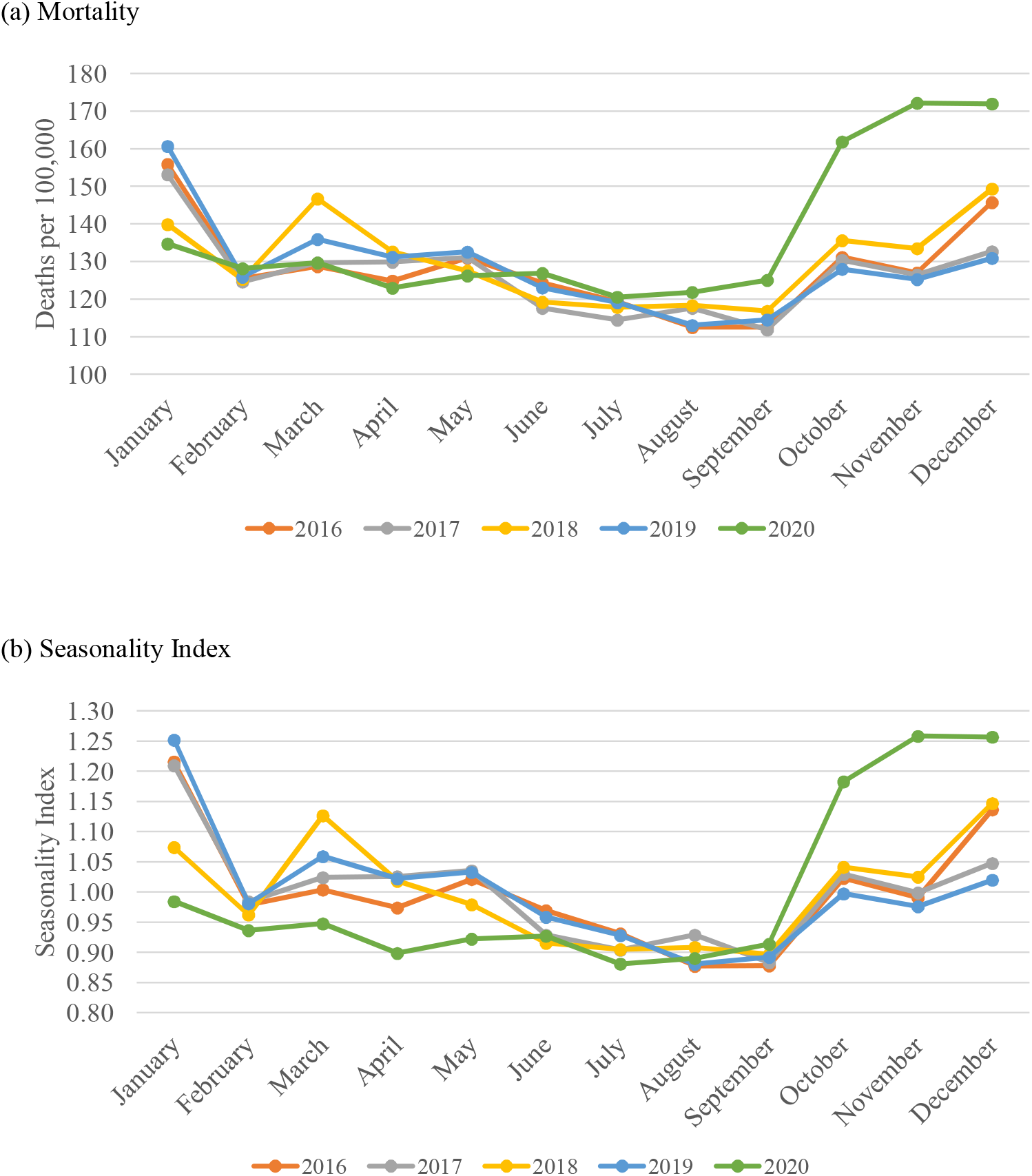
Mortality (deaths per 100,000 population) and seasonality index by calendar month and year in government-controlled Ukraine, all ages and both sexes combined Note: Population size as of January 1 of calendar year. Seasonality index is a ratio of the mortality in each month divided by the average monthly mortality.

Table 2 shows monthly case counts, recorded deaths, predicted deaths, and excess deaths. Ukraine recorded 1,091,490 COVID-19 cases during the calendar year. Positive values for excess deaths indicate that the number of 2020 recorded deaths exceeded the number of 2020 predicted deaths and negative values indicate that the number of recorded deaths was lower than that of the expected deaths. Overall, the model predicted 574,462 total deaths for the calendar year. There were 612,557 deaths recorded in 2020 indicating that 38,095 deaths or 6% were excess deaths for the entire year. In four calendar months (January, March, April, May) the number of predicted deaths exceeded that of recorded deaths resulting in negative excess death estimates. Deaths in January were 13% lower than expected and those in March and April were 6% and 7% lower, respectively. Excess deaths were positive in June and grew larger in each successive month thereafter peaking in November when excess deaths represented 26% of all deaths. If we consider only June to December the number of excess deaths were 52,124 or 14% of all deaths in those months.

**Table 2.**
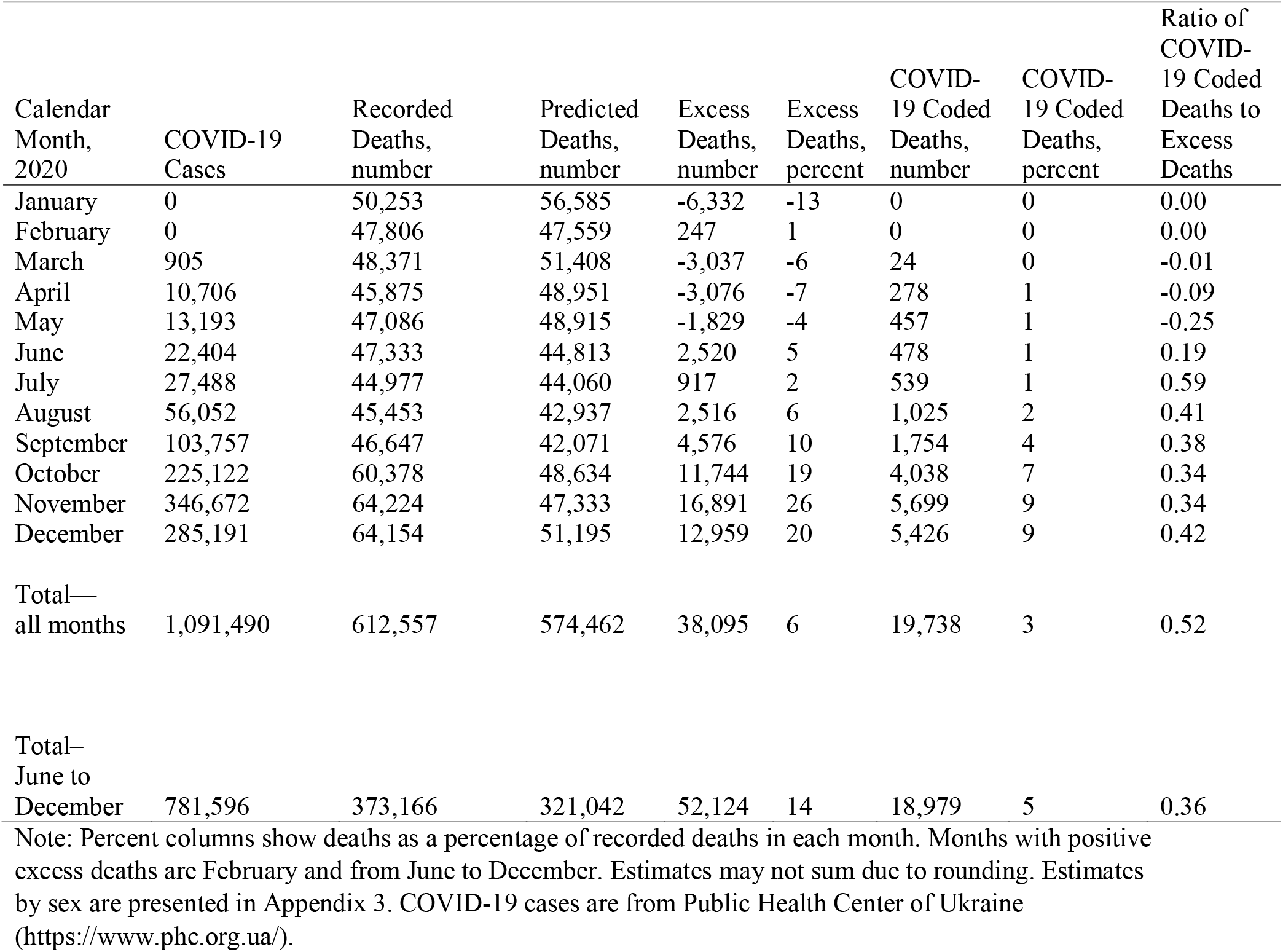
Recorded, predicted, and excess deaths in 2020 by calendar month; all ages and both sexes combined

Table 2 also shows the ratio, by month, of COVID-19 coded deaths with that of excess deaths. For the entire year, this ratio was 0.52 indicating that the total number of COVID-19 deaths was approximately one-half that of the total number of excess deaths. For June to December it was 0.36. Appendix 2 shows the 95% confidence intervals for the monthly excess death estimates.

Appendix 3 presents the sex stratified results. Excess death patterns were generally similar by sex although females had a higher estimated percentage of excess deaths compared to males—16% vs. 12%—during June to December. Appendix 3 shows that the ratio of COVID-19 coded deaths to excess deaths differed by sex. The ratio was 0.72 for males and 0.39 for females for the entire year. The differential was more modest—.45 for males and 0.30 for females— during June to December.

Age-specific estimates of excess deaths and COVID-19 coded deaths are presented in Figure 2 and Appendix 4. We limit the period to June to December when excess mortality was positive for all ages combined in each month. Figure 2 shows the proportion of all recorded deaths that were excess deaths and COVID-19 coded deaths, respectively. Appendix 4 provides the actual number of predicted and excess deaths by age groups. It is notable that positive excess mortality was not limited to those over age 65. All age groups over ages 35-39 experienced positive excess mortality with the oldest ages experiencing the largest proportionate increases. Those ages 70-74 experienced the highest proportionate increase in mortality at about 32% of recorded deaths. Among those under the age of 25, three age groups (0-4, 15-19, 20-24) experienced positive excess mortality. The absolute number of recorded deaths in the younger age groups was relatively small so although these groups experienced positive excess mortality the absolute number of excess deaths was also small. For example, among those aged 20-24 there were 938 deaths recorded from June to December and 821 deaths predicted (Appendix 4) resulting in a 12.4% excess mortality.

**Figure 2.**
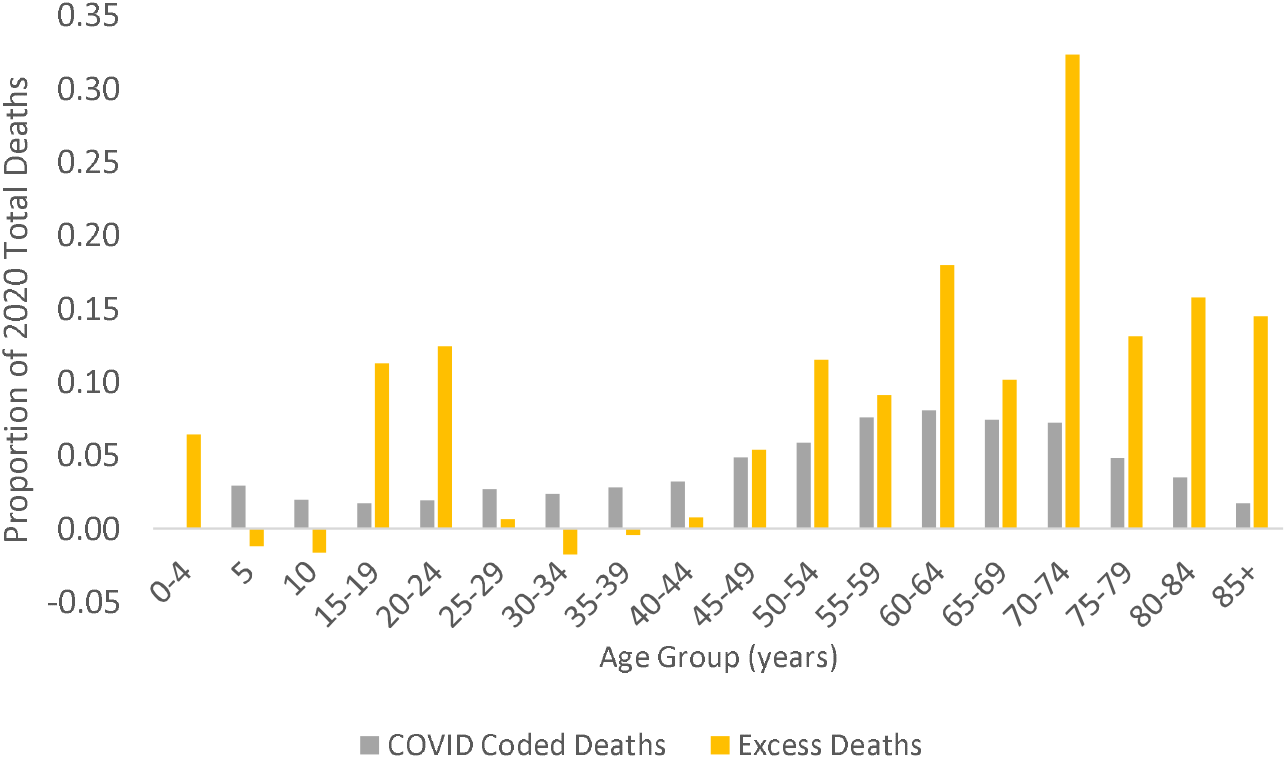
COVID-19 coded deaths and 2020 excess deaths by age group, June to December 2020

Figure 3 compares our estimates of excess deaths with estimates with 30 European countries published by Eurostat for EU countries. The estimates produced by Eurostat are calculated from the average number of monthly deaths in each country during 2016-2019 and thus differed from our methodology. Figure 1a compares the Ukrainian experience with the median and upper and lower quartiles of the 30 EU countries. Also included in the figure is the level of the country with the highest percentage of excess deaths in each month. Six countries experienced very high levels of excess deaths in April: Belgium, Spain, Netherlands, Italy, Sweden, and France. Ukraine did not experience this peak. From May onwards excess deaths for Ukraine stayed remarkably close to that of the median of the 30 EU countries. Figure 3b places Ukraine in the context of four European countries in close geographic proximity. Excess deaths in Ukraine again tended to be close to the average of the four countries beginning in May. Ukraine also avoided a large peak in the later part of the year in contrast to the four other countries.

**Figure 3.**
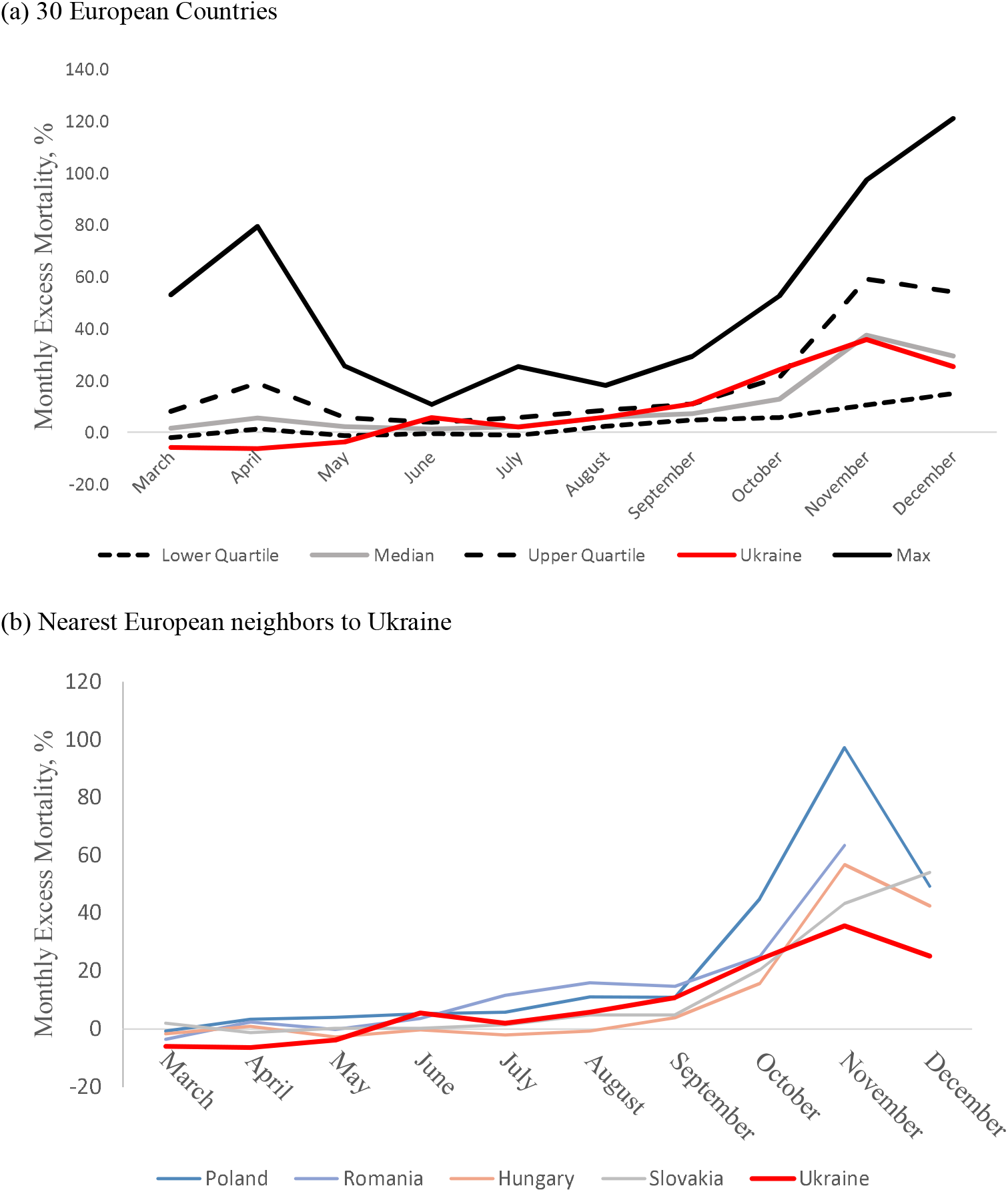
Comparison of excess deaths in Ukraine with the European distribution and nearest neighbors

Appendix 5 highlights how estimates of excess deaths based on the “average level” of death can be highly sensitive to the time-period used for the calculation in situations where there is rapid demographic change. Appendix 5 shows that estimates of excess mortality in Ukraine decline the longer the time-period used as a basis for the calculation. This pattern results because the number of annual deaths decreases over time and including earlier years in the average results in an underestimate of expected deaths in 2020.

## Discussion

Using a recent death trends, we estimated monthly excess deaths for Ukraine in 2020. The model resulted in an estimate of 38,095 excess deaths for 2020 and 52,124 excess deaths if we consider only June to December. Our results need to be interpreted in the context of the strengths and limitations of the excess death metric and our approach. The main strength of the metric is that it does not rely on the capacity of a country to detect all COVID-19 cases and code deaths resulting from coronavirus infection on the death certificate accordingly. The metric also provides a yardstick to measure the full death toll of the pandemic including deaths directly attributable to the virus and deaths attributable to the broader social and economic shocks that nations underwent. By incorporating recent trends in our model, rather than the “average” approach that is commonly used, we leveraged the stability of mortality trends. The “average” approach using 2015-2019 data would have resulted in an estimate of 26,200 excess deaths for 2020, smaller than our estimate. A main limitation of the excess death metric is that it does not isolate the directly attributable to coronavirus infection from other deaths that may be associated with social and economic upheavals created by the pandemic.

Ukraine’s aggressive action in terms of limiting social interactions in early March likely limited deaths during this early period. Unlike other European countries, Ukraine did not experience an early death surge. The country recorded its first confirmed COVID-19 coded deaths in March (24 deaths). Although 302 COVID-19 coded deaths were recorded in March and April combined, our estimates indicate that Ukraine experienced approximately 6,000 less deaths than would be expected in these two months. An additional 1,800 negative excess deaths were estimated for May. We can speculate that these “negative” estimates are partly attributable to a combination of lower mortality from influenza and reduced deaths from accidents and injuries resulting from restriction of movement. Increasing international evidence points to 2020 as having below average influenza activity as social distancing restrictions and severe limitations on international travel in response to the COVID-19 pandemic reduced global influenza spread [10]. Aside from reduced influenza, reduced deaths from traffic and other accidents and injuries associated with public interactions may also have been an important factor as has been shown in other countries [11]. Future studies examining causes of death will help identify the sources of the negative excess mortality.

Sustained and positive excess deaths were not observed until June soon after the easing of restrictions at the end of May. Similar to the growth in COVID-19 cases, excess deaths continued to grow at a steady pace reaching a peak in November. If we consider excess deaths beginning in June we estimated that 52,214 more individuals died than expected. In November, excess mortality was estimated to be 26% a level comparable to EU as a whole in April [2] but lower than peak estimates observed in the hardest hit countries. For example, the United Kingdom reached a peak excess mortality of 108% in the middle of April 2020 [3].

We found that COVID-19 coded deaths were approximately one-third that of our estimates of excess deaths for June to December. This finding is in line with studies from other countries that show that estimates of excess deaths often far exceed that of COVID-19 coded deaths [12,13]. The main conclusion is that reliance on COVID-19 coded deaths will substantially under-estimate the total mortality impact of the pandemic. One likely source for the discrepancy is that not all cases and deaths attributable to COVID-19 infection are detected because of insufficient testing. Another source is that excess deaths captures deaths directly attributable to infection as well as those indirectly caused by the social and economic upheavals of the pandemic.

Our estimates were based on linear models using data from the previous five years and as we show the model predicted well mortality in these earlier years with only a small degree of error. Our estimates of excess deaths are comparable in scope to those recently published in the Economist, which was also based on recent death trends and found negative excess mortality early in the year and growing positive excess mortality from June [14]. They estimated 30,710 excess deaths from March 31^st^ to November 29^th^, while we estimated 34,249 for a similar period (April-November).

An important finding emerging from our study is that the ratio of COVID-19 coded to excess deaths was lower in women compared to men—for June to December it was 0.30 in women and 0.45 in men. This finding is suggestive that women were less likely to have COVID-19 coded on the death certificate compared to men conditional on dying from the infection. The reasons for the gender discrepancy, however, is not clear and should be investigated in future work. Our estimates also highlight that excess mortality was not limited to older age groups, although these groups experienced the highest absolute increases in mortality. Particularly noteworthy is that those in age groups 0-4, 15-19, and 20-24 experienced positive excess mortality. While these groups may be at low risk of death from the virus itself, they may have been adversely affected by quarantine restrictions and effects on healthcare. For example, higher infant mortality may be related to poorer pre- and peri-natal care due to overburdened health systems or fear of accessing healthcare. For those in their teenage and early adulthood years, social isolation may have increased suicide risks. Domestic violence may have also increased along with other forms of violence. It is imperative that future studies elucidate the causes of excess mortality in these younger age groups.

In sum, we estimate that Ukraine experienced 38,095 excess deaths in 2020, which amounts to a 6% excess death rate. If we consider only June to December, the number of excess deaths was estimated to be 52,124 representing a 14% higher death rate for this period. Further studies are required to examine the causes of death that contribute to excess mortality during the year, the gender differences in COVID-19 coding on the death certificate, and the unusually higher than expected mortality experienced by certain younger aged groups. Also valuable will be sub-national estimates to identify whether positive excess mortality was experienced by certain regions or cities within Ukraine early in the pandemic.

## Data Availability

Requests for data and statistical code may be made to Olena Dorenshko.

## Acknowledgements

This publication has been made possible with the support of Switzerland. Specifically, this study was funded by the ‘Support to Reforms and Governance in the Health Sector’ project implemented by the World Bank and funded by the Swiss Agency for Development and Cooperation (SDC). The study was done by the World Bank Ukraine staff and consultants, who have also drafted the publication. The opinions and views presented in this publication represent those of the authors and may not necessarily represent the views of the World Bank, or the SDC. This work relies entirely on a secondary analysis of aggregated data without personal identifying information and therefore does not constitute human subjects research.

## Declarations

### Availability of data and material

Requests for data and statistical code may be made to Olena Dorenshko.

### Code availability

Requests for data and statistical code may be made to Olena Dorenshko.

### Authors’ contributions

All authors contributed to the scientific content of the manuscript. Neil Mehta wrote an initial draft and all authors edited the manuscript. Ihor Honchar conducted the statistical analyses.

### Ethics approval

This was a secondary data analysis of de-identified data and therefore does not constitute humans subjects research.

### Consent to participate

Not applicable.

### Consent for publication

All authors consent to publication.

## Supplementary Materials

**Appendix 1.**
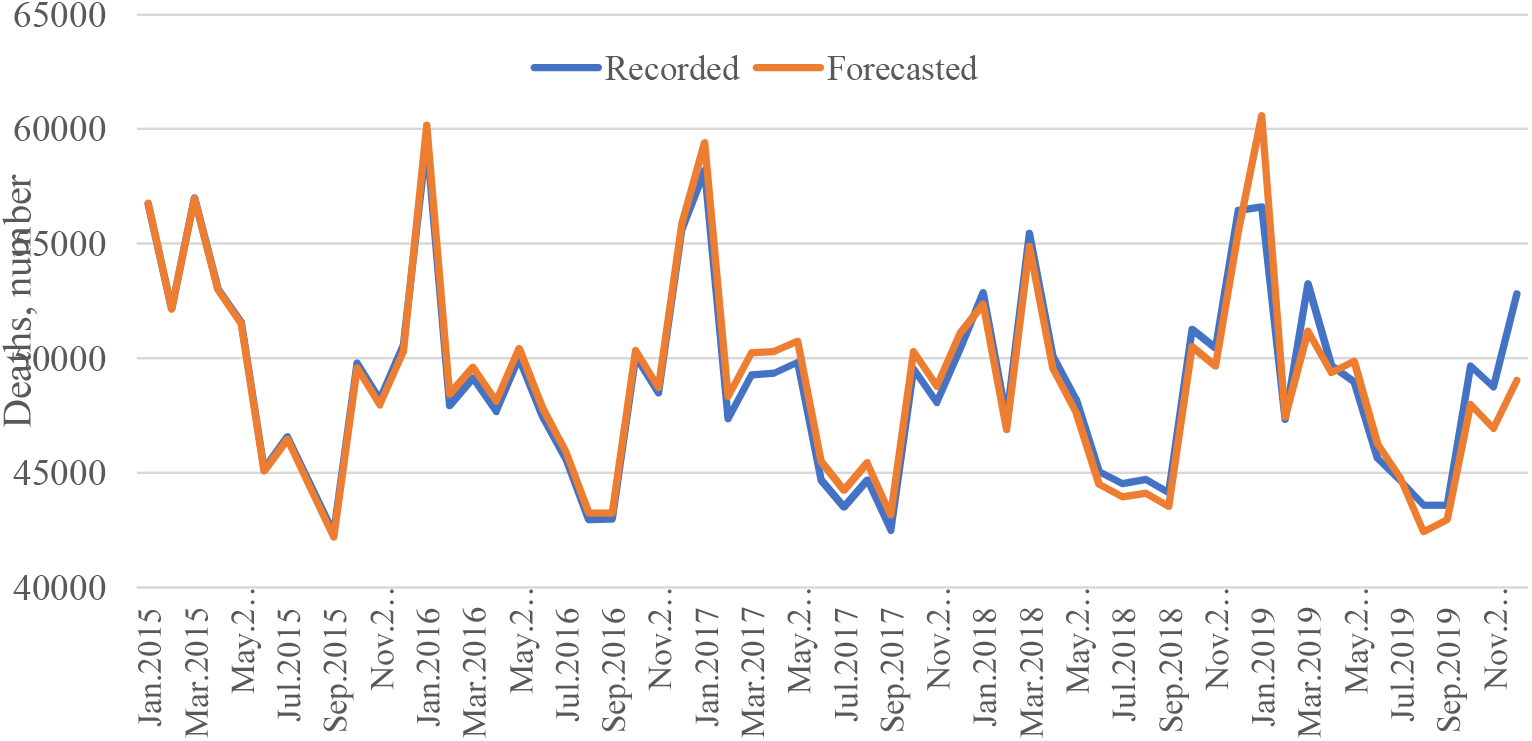
Model-based predicted and actual deaths by calendar month, 2015-2019 Note: Both sexes and all ages combined. Predicted deaths are based on the predictions from the regression model multiplied by the 2015-2019 average seasonal index for each month.

**Appendix 2.**
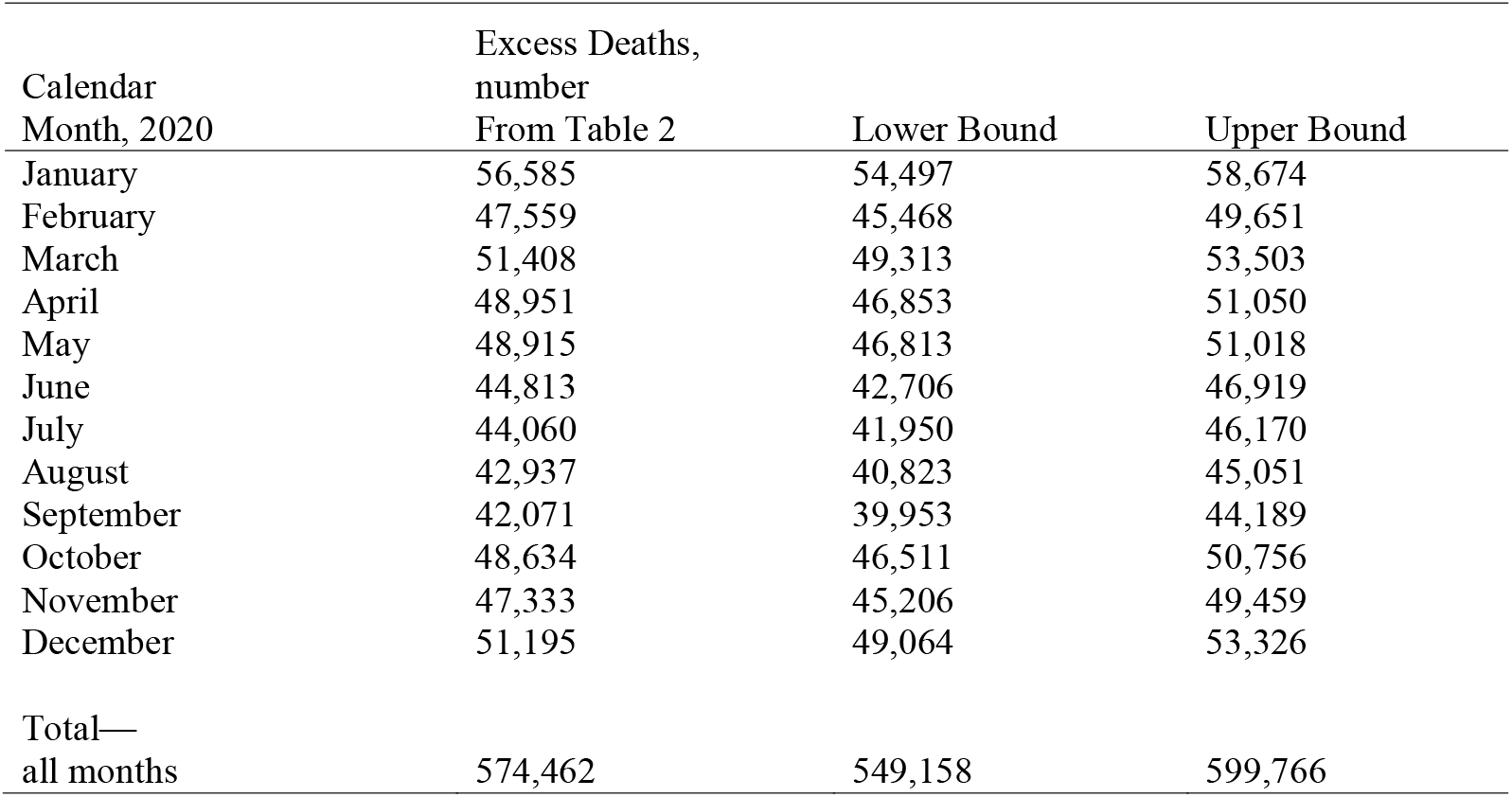
95% Confidence intervals for the monthly excess death estimates shown in Table 2.

| Calendar<br>Month, 2020 | Excess Deaths,<br>number<br>From Table 2 | Lower Bound | Upper Bound |
| --- | --- | --- | --- |
| January | 56,585 | 54,497 | 58,674 |
| February | 47,559 | 45,468 | 49,651 |
| March | 51,408 | 49,313 | 53,503 |
| April | 48,951 | 46,853 | 51,050 |
| May | 48,915 | 46,813 | 51,018 |
| June | 44,813 | 42,706 | 46,919 |
| July | 44,060 | 41,950 | 46,170 |
| August | 42,937 | 40,823 | 45,051 |
| September | 42,071 | 39,953 | 44,189 |
| October | 48,634 | 46,511 | 50,756 |
| November | 47,333 | 45,206 | 49,459 |
| December | 51,195 | 49,064 | 53,326 |
| Total—<br>all months | 574,462 | 549,158 | 599,766 |

**Appendix 3.** Recorded, predicted, and excess deaths in 2020 by calendar month and sex; all ages combined

| Males |  |  |  |  |  |  |  | Females |  |  |  |  |  |  |
| --- | --- | --- | --- | --- | --- | --- | --- | --- | --- | --- | --- | --- | --- | --- |
| Calendar Month | Recorded Deaths | Predicted Deaths | Excess Deaths, number | Excess Deaths, percent | COVID-19 Coded Deaths, number | COVID-19 Coded Deaths, percent | Ratio of COVID-19 Coded Deaths to Excess Deaths | Recorded Deaths | Predicted Deaths | Excess Deaths, number | Excess Deaths, percent | COVID-19 Coded Deaths, number | COVID-19 Coded Deaths, percent | Ratio of COVID-19 Coded Deaths to Excess Deaths |
| January | 25,038 | 28,042 | -3,004 | -12 | 0 | 0 | 0.00 | 25,215 | 28,544 | -3,329 | -13 | 0 | 0 | 0.00 |
| February | 23,196 | 23,394 | -198 | -1 | 0 | 0 | 0.00 | 24,610 | 24,166 | 444 | 2 | 0 | 0 | 0.00 |
| March | 23,878 | 25,474 | -1,596 | -7 | 7 | 0 | 0.00 | 24,493 | 25,934 | -1,441 | -6 | 17 | 0 | -0.01 |
| April | 22,685 | 24,483 | -1,798 | -8 | 166 | 1 | -0.09 | 23,190 | 24,468 | -1,278 | -6 | 112 | 0 | -0.09 |
| May | 23,207 | 24,521 | -1,314 | -6 | 252 | 1 | -0.19 | 23,879 | 24,394 | -515 | -2 | 205 | 1 | -0.40 |
| June | 23,399 | 22,627 | 772 | 3 | 229 | 1 | 0.30 | 23,934 | 22,185 | 1,749 | 7 | 250 | 1 | 0.14 |
| July | 22,549 | 22,402 | 147 | 1 | 296 | 1 | 2.01 | 22,428 | 21,658 | 770 | 3 | 243 | 1 | 0.32 |
| August | 22,794 | 21,906 | 888 | 4 | 567 | 2 | 0.64 | 22,659 | 21,030 | 1,629 | 7 | 458 | 2 | 0.28 |
| September | 23,149 | 21,529 | 1,620 | 7 | 935 | 4 | 0.58 | 23,498 | 20,542 | 2,956 | 13 | 819 | 3 | 0.28 |
| October | 30,096 | 24,799 | 5,297 | 18 | 2,115 | 7 | 0.40 | 30,282 | 23,835 | 6,447 | 21 | 1,925 | 6 | 0.30 |
| November | 31,804 | 23,994 | 7,810 | 25 | 3,048 | 10 | 0.39 | 32,420 | 23,339 | 9,081 | 28 | 2,650 | 8 | 0.29 |
| December | 31,661 | 25,729 | 5,932 | 19 | 2,883 | 9 | 0.48 | 32,493 | 25,466 | 7,027 | 22 | 2,561 | 8 | 0.36 |
| Total—<br>all months | 303,456 | 288,901 | 14,555 | 5 | 10,498 | 3 | 0.72 | 306,625 | 285,561 | 23,540 | 8 | 9,240 | 3 | 0.39 |
| Total—<br>June to<br>December | 185,452 | 162,987 | 22,465 | 12 | 10,073 | 5 | 0.45 | 187,714 | 158,055 | 29,659 | 16 | 8,906 | 5 | 0.30 |

Note: Percent columns show deaths as a percentage of recorded deaths. Estimates may not sum due to rounding.

**Appendix 4.** Recorded, predicted, and excess deaths from June to December 2020 by age group; both sexes combined

| Age Group (years) | Recorded Deaths, number | Predicted Deaths, number | Excess Deaths, number | COVID-19 Coded Deaths, number |
| --- | --- | --- | --- | --- |
| 0-4 | 1,409 | 1,318 | 90 | 0 |
| 5-9 | 207 | 209 | -2 | 6 |
| 10-14 | 255 | 259 | -4 | 5 |
| 15-19 | 579 | 514 | 65 | 10 |
| 20-24 | 938 | 821 | 117 | 18 |
| 25-29 | 1,860 | 1,849 | 11 | 50 |
| 30-34 | 4,261 | 4,336 | -76 | 101 |
| 35-39 | 6,309 | 6,338 | -29 | 177 |
| 40-44 | 8,680 | 8,615 | 64 | 276 |
| 45-49 | 11,236 | 10,635 | 601 | 545 |
| 50-54 | 14,442 | 12,779 | 1,663 | 842 |
| 55-59 | 22,739 | 20,673 | 2,066 | 1,719 |
| 60-64 | 33,147 | 27,197 | 5,950 | 2,664 |
| 65-69 | 40,485 | 36,382 | 4,102 | 3,005 |
| 70-74 | 44,440 | 30,075 | 14,364 | 3,204 |
| 75-79 | 45,568 | 39,599 | 5,968 | 2,184 |
| 80-84 | 70,262 | 59,195 | 11,067 | 2,428 |
| 85+ | 65,376 | 55,927 | 9,448 | 1,119 |
| Total | 372,187 | 316,721 | 55,466 | 18,353 |

Note: Predicted deaths based on age-specific regression models. Excludes deaths of unknown age. Estimates may not sum due to rounding.

**Appendix 5.** Comparison of estimates of excess deaths in Ukraine for 2020 based on using the average number of deaths across various time periods

| Time Interval | Average Annual Number of Deaths in Interval (thousands) | 2020 Excess Mortality (thousands) | Percentage Higher than 2020 COVID-19 Coded Deaths, % |
| --- | --- | --- | --- |
| **2015-2019** | 586 | 26.2 | 34 |
| **2016-2019** | 583 | 29.1 | 48 |
| **2017-2019** | 582 | 30.4 | 55 |
| **2018-2019** | 585 | 28.0 | 43 |
| **2019** | 578 | 34.2 | 74 |

Note: COVID-19 Coded deaths

